# Improving HIV disclosure to sexual partners via assisted partner notification services: A mixed-method study on utilization, barriers, and facilitators

**DOI:** 10.1101/2025.11.06.25339738

**Authors:** Alex Gabagambi Alexander, Blandina T. Mmbaga, Edna Paul, Noela H. Daniel, Mariam L. Barabara, Aloyce M. Mlyomi, Florida J. Muro, John A. Barlett, Charles Muiruri

## Abstract

**Background:** The global burden of HIV remains substantial, with Tanzania among the most affected countries in sub-Saharan Africa. The central regions of Tanzania such as Singida region, has high new HIV infections, partly driven by low voluntary HIV disclosure among couples. Assisted Partner Notification Services (APNS) have emerged as a promising strategy to support HIV disclosure, promote partner testing, and reduce transmission. Despite this potential, APNS remains underutilized due to a range of individual, social, and structural barriers.

**Objectives:** This study aimed to assess the utilization of APNS and identify barriers and facilitators influencing its uptake among people living with HIV (PLWH) in Singida, Tanzania.

**Methods:** We employed a convergent mixed-methods design. A cross-sectional survey quantified APNS utilization and associated factors among PLWH with sexual partners, while qualitative interviews explored barriers and facilitators from the perspectives of both PLWH and healthcare providers (HCPs).

**Results:** Only 40% of participants reported APNS utilization. Higher knowledge of HIV disclosure was positively associated with uptake (AOR = 2.65, 95% CI: 2.28–2.81; p = 0.02), whereas depressive symptoms reduced engagement (AOR = 0.95, 95% CI: 0.91– 0.99; p = 0.027). Awareness of pre-exposure prophylaxis (PrEP) was strikingly low (13%). Reported barriers included stigma, fear of financial and emotional insecurity, cultural constraints, and weak health system support. Facilitators involved emotional readiness, trust in providers, mobile outreach, and supportive community structures. Participants recommended enhanced APNS education, mental health integration, strengthened provider training, and improved access through transport and outreach interventions.

**Conclusion:** Findings reveal low APNS uptake in Singida, constrained by stigma, mental health challenges, and systemic gaps. Strengthening disclosure knowledge, provider trust, and outreach services, while integrating APNS with PrEP and chronic disease care, may enhance impact and reduce HIV transmission.

## Introduction

The global burden of HIV remains substantial. According to the UNAIDS 2023 report, 39 million people worldwide were living with HIV, with 1.3 million newly diagnosed in 2022 [1]. Notably, 4.4% of these individuals (1.7 million) were from Tanzania, where 2.5% (32,000) of all newly diagnosed cases globally also occurred [1]. In Central Tanzania, the Singida region has experienced a persistent rise in new HIV infections over the past decade compared to other regions. The Tanzanian National AIDS Control Program (NACP) report from 2020 indicated that Singida had an HIV prevalence rate of 3.6% and a high rate of new HIV infections [2, 3]. This trend is primarily driven by heterosexual transmission among couples, a group historically characterized by low rates of voluntary HIV disclosure [4]. Several studies have documented stagnant rates of voluntary HIV disclosure in this region [2, 5, 6]. A recent study by Alexander et al. (2024) in the study area found that approximately 22% of clients with sexual partners attending Care and Treatment Centers (CTCs) had not voluntarily disclosed their HIV status to their partners [7]. This significant proportion of non-disclosure highlights the need for greater emphasis on assisted partner notification services (APNS), which provide structured support for people living with HIV (PLWH) to disclose their HIV status to sexual partners [7]. However, despite its potential, there are limited studies in Tanzania assessing the utilization of APNS and the barriers and facilitators influencing its uptake [8].

APNS is a public health strategy in which a trained service provider assists consenting PLWH in notifying their sexual partners of potential exposure to HIV [9]. It is a valuable HIV prevention tool that promotes voluntary HIV disclosure to sexual partners [10]. APNS can be conducted using three main approaches: passive referral, where the HIV index client independently notifies their partner(s); contract referral, where the client agrees to notify their partner within a specified period before the provider follows up; and provider referral, where a health worker directly notifies the partner while maintaining the client’s confidentiality [11]. In most areas, passive and provider referral are the main strategies used. For example, Mugisha et al.(2023) revealed that 43% of providers using APNS reported using the passive referral strategy, and 41% reported using the provider referral [12].

Evidence shows that APNS significantly improves partner testing and linkage to care. One of the studies revealed that disclosure with APNS resulted in a higher number of partners tested and better linkage to treatment for HIV-positive partners by 1.5 fold compared to those who had not used APNS [13, 14]. Strong evidence supports APNS as a critical method for reaching at-HIV risk populations and contributing to the UNAIDS 95-95-95 targets [15]. The studies conducted in sub-Saharan Africa found that APNS is feasible and effective in improving HIV case-finding [15, 16]. APNS was a prominent feature of issued World Health Organization (WHO) guidance on couples’ HIV testing and counseling, including antiretroviral therapy (ART) for treatment and prevention in sero-discordant couples [17]. It is also recommended that APNS be done voluntarily, and all sexual partners of PLWH should be offered APNS [17, 18]. Despite this, the uptake of APNS remains inconsistent. The utilization of APNS is reported at 69% in Kenya, 65.5% in Ethiopia, 56% in Eastern Tanzania, and 62% in Northern Tanzania [15, 18–20]. Tanzanian national guidelines on HIV testing encourage the use of APNS; however, despite the favorable policy environment to use this intervention in this area, its adoption is limited among PLWH in Tanzania [21].

Despite its proven effectiveness, the utilization of APNS remains limited, particularly among men, youth, and rural populations [22]. Studies indicate that these populations are less likely to participate in APNS due to a variety of factors, including individual, interpersonal, healthcare system, and societal factors [9, 12, 15, 18, 22–24]. Moreover, several studies have highlighted a range of barriers that hinder the utilization of APNS and HIV disclosure among PLWH, including stigma, embarrassment, fear of rejection or violence, lack of confidentiality, lack of knowledge about APNS, a lack of trust in the HIV Testing and Counselling (HTC) counselor, unwillingness to notify partners, unfriendly services, and poor provider-client rapport [9, 25–27]. These challenges are compounded by psychological stress, cultural and educational differences, and societal discrimination, particularly among key populations [28, 29]. Conversely, facilitators that enhance APNS uptake include effective provider communication, personal moral responsibility, relational intimacy, partner understanding, professional support, peer encouragement, and improved public HIV literacy [29, 30]. Together, these findings reveal a complex interplay of influences shaping APNS engagement.

This study aims to assess the utilization of APNS, its associated factors, barriers, and facilitators among PLWH with sexual partners to improve HIV disclosure in the Singida region. We hypothesize that APNS supports HIV disclosure to partners [32], and that its use is shaped by socio-cultural and structural factors such as stigma, mental health, and systemic barriers, while enablers include knowledge, provider trust, and community outreach, findings that will guide tailored strategies to improve policy, service delivery, and HIV outcomes in high-burden areas

## Materials and methods

### Overall study design

We employed a convergent mixed-methods design in this study to capture both the measurable patterns of APNS service uptake and the deeper contextual factors influencing its use among PLWH and HCPs. This approach enabled triangulation of findings and a richer understanding of the complexities surrounding APNS utilization. Quantitative data highlighted key trends in APNS uptake and demographic associations, while qualitative data provided insight into experiences, motivations, fears, and systemic barriers.

### Conceptual framework

We employed the **Social Ecological Model (SEM),** which provides a comprehensive framework for understanding the utilization and factors that influence the use of APNS. The SEM emphasizes that health behaviors are shaped by multiple, interacting levels of influence: individual, interpersonal, community, and societal [29, 31]. It has been previously applied to analyze APNS among men who have sex with men (MSM), but has not yet been systematically used to study APNS among heterosexual PLWH in Tanzania [29]. Moreover, the framework can also be used to summarize the facilitators and barriers to using APNS. This was the first study to use the SEM to systematically study APNS among PLWH with sexual partners (Fig 1).

**Fig 1:**
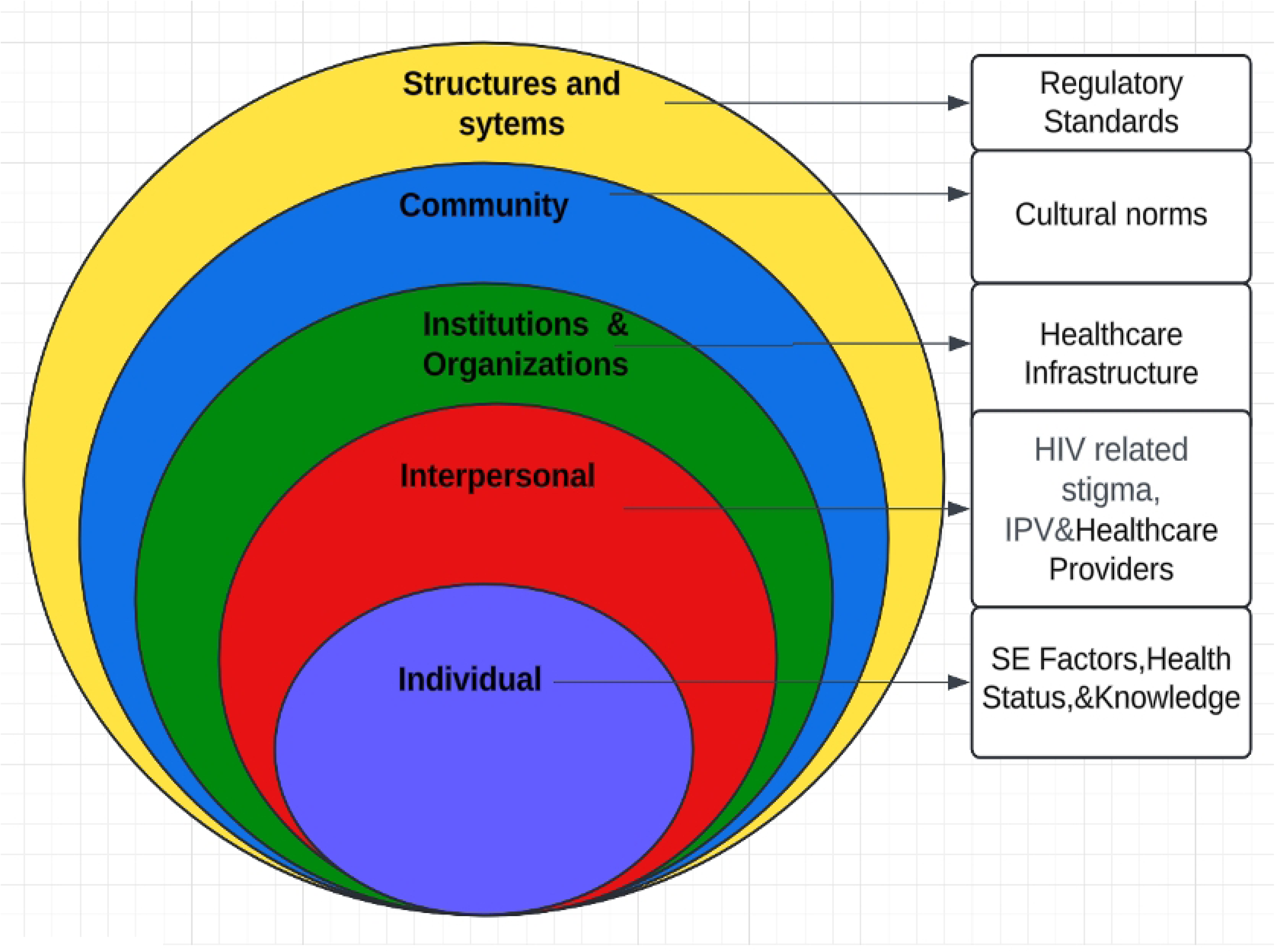
Illustration of the Social Ecological Model (SEM)

### Quantitative strand

#### Study design and study area

This was a hospital-based cross-sectional study. Data was collected between June-July 2025 at Singida Regional Referral Hospital (SRRH) in the Singida region of the United Republic of Tanzania.

#### Study population and sampling

Non-probability convenience sampling was used to recruit PLWH aged 18 years old and above, who had sexual partners since when they were diagnosed with HIV and who are linked to care and treatment (CTC) services for at least three months [33]. PLWH who mutually tested for HIV with his or her sexual partner were excluded. The sample size was calculated using the Kish and Leslie formula size estimation and sampling technique to determine the utilization of APNS for a single proportion N = Z2α/2(1−p) d2 [33], referring to a study conducted in Eastern Tanzania [20], where Z-alpha is the standard normal value corresponding to the set confidence level of 95% = 1.96, d is the tolerable sampling error (precision) = 5%, p is the estimated prevalence of APNS utilization in the study = 0.62 [15] and N is the required sample size = 300. With an assumed 10% non-response rate, the total sample size required was 330, and we had a total of 337 participants.

#### Data collection

Trained research assistants (RAs) who were proficient in Swahili, and staff members at the CTC clinic collected data using a structured and pre-tested interviewer-administered questionnaire. To reduce social desirability bias, female participants were interviewed by a female interviewer and male participants by a male interviewer. The questionnaire was first prepared in English, and then translated into Swahili, and then translated back to English to check for consistency of the questions. The final version of the questionnaire was reviewed by a three-member expert panel (HIV disease and HIV-related stigma experts) to establish content validity.

Study participants were identified in terms of the eligibility by conducting screening interviews when individuals came for a scheduled CTC appointment. the study RAs approached the individual before their scheduled appointments and provided information about the purpose of the study. Those who consented to participate were provided with written informed consent and were interviewed after completing the CTC appointment activities. For participants who could not read or write, a hand-signature method mark on the document was applied to indicate their informed consent. The adapted and structured survey, which took approximately one hour to complete, was administered [18]. Enrolled participants completed an interviewer-administered survey capturing socio-demographic information, the experience of PLWH in APNS utilization, psychological factors (depression), and health system factors related to partner notification for HIV testing [18]. The SEM was also employed for the selection of significant variables when adapting the survey guide.

#### Measures

When we assessed the outcome variable, participants were asked if they had participated in APNS. Those who responded “yes” were coded as having participated, and those who responded “no” were coded as not having participated. The demographic variables included age, gender, highest education level, marital status, religion, and area of residence. The psychological factors included HIV-related stigma and depression. The health system factors related to partner notification, including ART medication adherence and blood pressure, were measured twice at different times, and the readings were used to determine hypertension status. A participant was classified as hypertensive if they had a systolic blood pressure of ≥140 mmHg and/or a diastolic blood pressure of ≥90 mmHg. We contextually adapted the short version (HSS-12) of the Berger HSS14 [34] to assess patient-perceived HIV-related stigma under four dimensions: (1) personalized stigma; (2) disclosure concerns; (3) negative self-image, and (4) concerns with public attitudes, each comprising a subscale of the instrument. 12 Items on this scale are rated from 1 to 4, with (1) being ‘strongly disagree’ and (4) ‘strongly agree. The total score ranges between 12 and 48 and is derived from the summation of item scores. Higher scores designate a greater level of perceived HIV-related stigma and the questionnaire has an internal reliability of 0.88 [34]. We have decided to use this short version because there is a wish to include a brief stigma component in our longer surveys to investigate the life situation of PLWH, in clinical contexts as a brief screening measure for signs of HIV stigma-related problems [35].

We also assessed knowledge of HIV disclosure through six questions. The maximum score for those questions was six, and those participants with ≥ five scores had high knowledge, four to three had moderate knowledge, and those with scores below three had low knowledge of disclosure [36].

The self-reported AIDS Clinical Trials Group (ACTG) adherence questionnaire was used to assess medication adherence. Adherence was treated as a continuous variable, and three questions from this questionnaire were used; and the score for adherence ranges from (0-100)% [37]. To know how many individuals were adherent to ART medications, the data that was obtained from this section on adherence from the ACTG was dichotomized into adherent > 90% of a total score and nonadherent when the total score was ≤ 90% [37, 38]. Additionally, we examined the most recent viral load of the client after being linked in APNS by recording it from the client’s CTC card. The client was considered to have a suppressed viral load count if he/she had HIV-1 RNA copies less than 50 copies/mL after disclosure [39].

We used a validated WHO-5 tool to assess depression. Its scores correlated well with those of the PHQ-9, suggesting that the WHO-5 represents a valid screening tool for depression. Nolan et al (2018) study revealed that the WHO-5 represents a useful tool in the Tanzania context and potentially in East Africa. Its rate of positive depression screens is high among PLWH in Tanzania [40]. The raw score is calculated by totaling the figures of the five answers. The raw score ranges from 0 to 25, with 0 representing the worst possible depression and 25 representing no depression. To obtain a percentage score ranging from 0 to 100, the raw score is multiplied by 4. A percentage score below 50 (or a raw score below 13) has been suggested as a cut-off for poor mental well-being and as an indication for further assessment of the possible presence of a mental health condition.

### Data analysis

Descriptive statistics were used to characterize the participants of our study. We planned to use multiple imputation to replace missing values before analysis. We conducted separate bivariate and multivariate logistic regression analyses to examine associations between participation in APNS and demographic characteristics. Notably, the factors in the regression model were selected based on SEM and published literature. For both time points, factors with a p-value of .25 or less in preliminary univariable analyses were included in final multivariable logistic regression models [41]. A final multivariable model was developed through backward stepwise elimination, retaining predictors significant at p<0.05. Multicollinearity was assessed using the Variance Inflation Factor (VIF). All independent variables had VIF values below 5, indicating that multicollinearity was not a concern in the final model. Variables that were statistically insignificant at the crude odds ratio (COR) level were not reported in the adjusted model, due to lack of significance in multivariable analysis. Data was analyzed using IBM SPSS for Mac, version 27.0. The SEM framework guided the reporting of our results. A significance level of α=0.05 was set for all tests to determine statistical significance.

### Qualitative strand

#### Study design

We employed a phenomenological qualitative design to explore the barriers and facilitators to APNS among PLWH with sexual partners, as well as the perspectives of HCPs. This approach allowed for an in-depth understanding of participants’ lived experiences with APNS. Using a semi-structured interview guide developed through literature review and expert input [25, 27, 29, 32], we purposively sampled participants with varied experiences of APNS, both those who had used the service and those who had not, ensuring diversity in age and sex among PLWH, and including HCPs (CTC staff) involved in delivering APNS services. We used a SEM to inform our analytical approach, recognizing that APNS uptake and implementation are influenced by multiple intersecting levels, ranging from personal and interpersonal factors to broader community norms, healthcare systems, and legal environments [29].

#### Data collection procedures

Data were collected through a combination of in-depth interviews (IDIs) and focus group discussions (FGDs). Among PLWH, we conducted 10 IDIs and five FGDs (each with five participants), while among HCPs, we conducted seven IDIs until thematic saturation was reached. All data collection activities were conducted in Swahili by trained RAs experienced in qualitative methods. Interviews and discussions took place in private rooms at the clinic to ensure confidentiality and comfort, and participants were given clear information about the study before providing informed consent. Sessions were audio-recorded and supplemented by detailed field notes.

The interview guide was piloted with a small number of eligible participants not included in the final analysis. Feedback from the pilot was used to improve question clarity, refine wording, and adjust topic sequencing to improve the conversational flow. Interviews and discussions lasted between 60 and 90 minutes. Audio recordings were transcribed verbatim in Swahili before being translated into English for analysis.

We employed data triangulation by drawing on multiple methods, including quantitative survey data with qualitative IDIs and FGDs, to explore consistent and divergent views on APNS. In addition, participant triangulation was applied by incorporating the perspectives of two distinct groups: PLWH and HCPs. This dual approach allowed for a more comprehensive and credible understanding of the APNS context from both the client and HCPs’ perspectives.

#### Researcher characteristics and reflexivity

The project had a multidisciplinary team that comprised researchers with backgrounds in public health, social sciences, and HIV program implementation. Several of the Tanzanian researchers had prior experience working with CTCs in the Singida region, which facilitated rapport with participants. At the same time, this insider position may have introduced assumptions about HIV disclosure norms in the region. To address this, we engaged in regular reflexive discussions and debriefings to question our presuppositions and interpretations. International team members, who were less embedded in the local health system, contributed to balancing potential bias by bringing comparative perspectives. We recognize that our prior belief in the potential of APNS may have shaped our inquiry, particularly in framing APNS as a critical intervention, and we attempted to mitigate this by grounding our analysis in participants’ accounts rather than preconceptions.

#### Data analysis

We conducted thematic analysis, supported by NVivo 12. The process began with a single team member summarizing and coding each interview transcript. Following the initial coding, a second team member reviewed all transcripts to ensure accuracy and clarity, as well as to evaluate the coding. This collaborative approach aims to enhance the reliability of the coding process and ensure a comprehensive understanding of the data.

After the review, both team members engaged in a consensus-building discussion to finalize the coding within each transcript. The next step involves combining qualitative memos and cross-analyzing them to identify core themes that emerge across the interviews. Representative quotes were then selected to provide compelling evidence for the synthesized results, ensuring that the findings reflect the perspectives of the participants while capturing the richness of the qualitative data. This systematic approach allowed for a robust analysis of the data, leading to meaningful insights into the research questions.

### Ethical consideration

The study was approved by KCMC University ethical committee (No. 2664) and the National Institute for Medical Research (NIMR) (No. NIMR/HQ/R.8a/Vol.IX/4866). Written informed consent was obtained from the study respondents individually. The right of the respondent to withdraw from the interview or not to participate at all was assured. To maintain participant confidentiality, the interviews were conducted in a private room with the door closed, and all of the documents with participants’ information were well protected in a locked cabinet; only the principal investigator had access to the key.

## Results

### Quantitative strand

A total of 337 respondents were enrolled in this study, with a 100% response rate. No participants had missing data for each variable of interest. The median age of participants was 50 years. Most participants were female, 258 (76.6%). More than half of the participants, 190 (56.4%) reported being in casual sexual relationships. Educational attainment was low among participants, with 249(73.9%) having completed only primary education. Out of all participants, 134 (40%) reported using APNS to disclose their HIV status to sexual partners. The majority of those who received APNS, 107 (79.5%) participants, accessed the service at a health facility. Among participants who used APNS, 108 (80.7%) reported knowing the HIV status of their sexual partner after disclosure. Of these, 50 (37.5%) sexual partners tested HIV-negative, while 58 (43.2%) sexual partners were HIV-positive status. Most participants, 290 (86.1%) of participants reported having never heard of PrEP before this study.

The participants who reported to have a chronic illness were 21(6.2%). Among PLWH who reported having a chronic illness, 11 individuals (52.4%) had hypertension, and 3 (14.3%) had diabetes mellitus. However, clinical assessments of the blood pressure conducted during the study revealed that 81 participants (23.8%) were hypertensive. Surprisingly, only 11(13.6%) participants among those whose BP was measured and found to be hypertensive were aware that they had hypertension and approximately 86.4% of hypertensive participants had undiagnosed hypertension (Table 1).

**Table 1:** Socio-Demographic and Clinical Related Characteristics of the Study Population(N=337)

| Variable | Category | Frequency (n) | Percent (%) |
| --- | --- | --- | --- |
| Age | 18-25 | 11 | 3.3 |
|  | 26-35 | 59 | 17.5 |
|  | 36-45 | 61 | 18.1 |
|  | Above 45 years | 206 | 61.1 |
| Gender | Male | 79 | 23.4 |
|  | Female | 258 | 76.6 |
| Marital status | Single | 69 | 20.5 |
|  | Married/ cohabiting | 111 | 32.9 |
|  | Divorced/Separated | 70 | 20.8 |
|  | Widowed | 87 | 25.8 |
| Type of sexual relationship | Primary partner | 147 | 43.6 |
|  | Casual partner | 190 | 56.4 |
| Residence | Rural | 59 | 17.5 |
|  | Urban | 278 | 82.5 |
| Occupation | Self employed | 231 | 68.5 |
|  | Employed | 20 | 5.9 |
|  | Unemployed | 86 | 25.5 |
| Religion | Muslim | 203 | 60.2 |
|  | Roman catholic | 94 | 27.9 |
|  | Protestant | 21 | 6.2 |
|  | Sabbath | 19 | 5.6 |
| Highest level of education | No formal education | 38 | 11.3 |
|  | Primary level | 249 | 73.9 |
|  | Secondary level | 44 | 13.1 |
|  | College/University level | 6 | 1.8 |
| Current chronic illness apart from HIV | Yes | 21 | 6.2 |
|  | No | 316 | 93.8 |
| Body mass index | Underweight | 38 | 11.3 |
|  | Normal weight | 193 | 57.3 |
|  | Overweight | 69 | 20.5 |
|  | Obese | 37 | 11.0 |
| Hypertension (from BP measurement) | Normotensive | 256 | 76.2 |
|  | Hypertensive | 81 | 23.8 |
| Duration on ART medication (Month) | Mean ±SD | 132.26±75 | |
| Viral Load | Viral load detected | 26 | 7.7 |
|  | Target not detected | 311 | 92.3 |
| CD4 count | Less 500 | 137 | 40.7 |
|  | Above 500 | 200 | 59.3 |
| Ever heard of PrEP before | No | 290 | 86.1 |
|  | Yes | 47 | 13.9 |
| Medication Adherence | Adherent > 90% | 251 | 74.5 |
|  | Nonadherent | 86 | 25.5 |
| Knowledge of HIV Disclosure | High knowledge | 255 | 75.7 |
|  | Moderate knowledge | 51 | 15.1 |
|  | Low knowledge | 31 | 9.2 |
| Depression | Depressed | 262 | 77.7 |
|  | Not Depressed | 75 | 22.3 |
| Anxiety | No/mild | 185 | 54.9 |
|  | Moderate/ severe | 152 | 45.1 |
| Perceived HIV Related stigma | Mean ±(SD) | 25.3±5.6 | |

### Types of APNS

This distribution indicates that nearly half of the APNS, 62 (46%), fall into the Passive category, while Provider and Dual APNS also represent substantial proportions. Contract APNS are very rare in comparison (Fig. 2).

**Fig 2:**
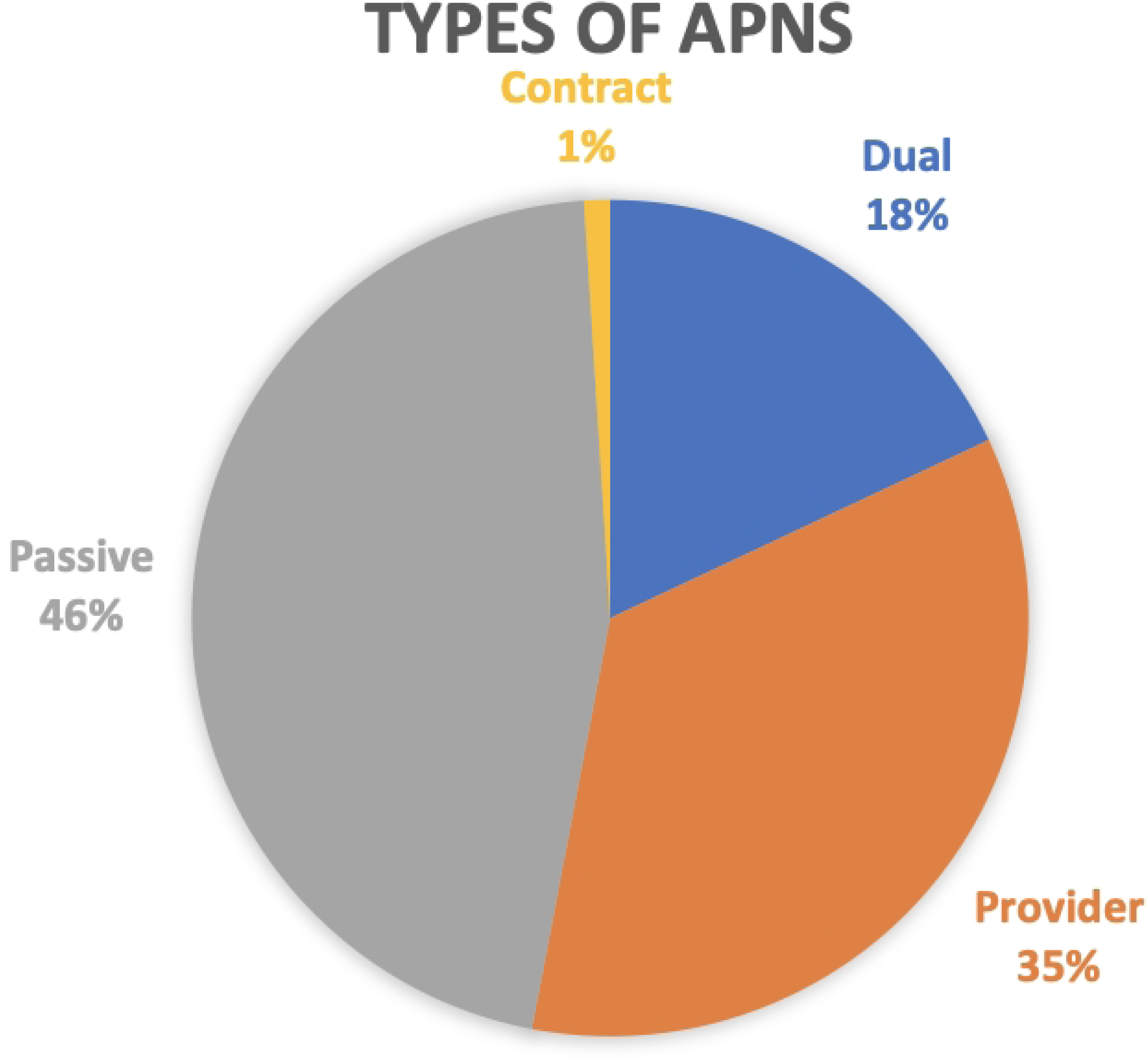
Pie chart showing the distribution of types of APNS.

### Factors associated with APNS Utilization

In the bivariate analysis, male participants had significantly higher odds of utilizing APNS compared to females (Crude OR = 1.81, 95% CI: 1.05–3.12; p = 0.032). Marital status was also significantly associated, with those who were married or cohabiting showing higher odds compared to those who had never married (Crude OR = 2.78, 95% CI: 1.38– 5.59; p = 0.004). Regarding the type of sexual relationship, individuals with casual partners had significantly lower odds of the utilization of APNS by 45% compared to those with primary partners (Crude OR = 0.55, 95% CI: 0.34–0.89; p = 0.02).

In the multivariate model, participants with moderate knowledge on HIV disclosure had significantly higher odds of utilizing APNS compared to those with low knowledge on HIV disclosure (Adjusted OR = 1.94, 95% CI: 1.06–3.58). Participants with high knowledge of HIV disclosure had 2.65 higher odds of utilizing APNS compared to those with low knowledge when keeping other factors constant (Adjusted OR = 2.65, 95% CI: 2.28–2.81). Lastly, higher depression levels were associated with reduced odds in adjusted models; each unit increase in depression score was associated with a 5% decrease in the likelihood of utilizing APNS when keeping other factors constant (Adjusted OR = 0.95, 95% CI: 0.91–0.99; p = 0.027) (Table 2).

**Table 2:** Bivariate and multivariate analysis; factors associated with utilization of APNS.

| Variable | Category | COR (95%CI) | P-value | AOR (95% CI) | P value |
| --- | --- | --- | --- | --- | --- |
| Gender | Female | 1 |  |  |  |
|  | Male | 1.81(1.05-3.12) | 0.032* |  |  |
| Marital status | Never married | 1 |  |  |  |
|  | Married or cohabiting | 2.78 (1.38-5.59) | 0.004* |  |  |
|  | Separated/Divorced | 0.81 (0.35-1.91) | 0.64 |  |  |
|  | Widowed | 0.89 (0.398-1.97) | 0.77 |  |  |
| Type of sexual relationship | Primary partner | 1 |  |  |  |
|  | Casual partner | 0.55 (0.34-0.89) | 0.02* |  |  |
| Ever heard of PrEP before | No | 1 |  |  |  |
|  | Yes | 1.75 (0.91-3.36) | 0.09 |  |  |
| Knowledge on HIV disclosure | Low knowledge | 1 |  | 1 |  |
|  | Moderate knowledge | 0.98 (0.5-1.92) | 0.95 | 1.94(1.06-3.58) | 0.03* |
|  | High knowledge | 2.28 (2.08-2.94) | 0.04* | 2.65(2.28-2.81) | 0.02* |
| Duration on ART medication |  | 1.004(1.001-1.008) | 0.011* |  |  |
| Depression |  | 0.94(0.89-0.99) | 0.02* | 0.95(0.91-0.99) | 0.027* |
| Note: *P-Value <0.05 |  |  |  |  |  |

### The barriers to APNS utilization

Among the participants, the most commonly reported barrier to APNS was fear of stigma, cited by 97(47.8%) of respondents. Fear of embarrassment 62 (30.4%) and fear of losing autonomy or emotional support 57 (28.3%) were also notable concerns. A smaller proportion of participants reported unwillingness to notify a partner 31 (15.2%), concerns about confidentiality 26 (13%), denial of HIV status 9 (4.3%), and not knowing their partner 4 (2.2%) as barriers (Table 3).

**Table 3:** Barriers to utilization of APNS(N=203)

| Characteristics | Category | Frequency | Percent (%) |
| --- | --- | --- | --- |
| Fear of Embarrassment/IPV | Yes | 62 | 30.4 |
|  | No | 141 | 69.6 |
| Fear autonomy and emotional support loss | Yes | 57 | 28.3 |
|  | No | 146 | 71.7 |
| Fear of stigma | Yes | 97 | 47.8 |
|  | No | 106 | 52.2 |
| Not knowing a partner | Yes | 4 | 2.2 |
|  | No | 199 | 97.8 |
| Unwillingness of a partner notification of partner. | Yes | 31 | 15.2 |
|  | No | 172 | 84.8 |
| Confidentiality | Yes | 26 | 13 |
|  | No | 177 | 87 |
| Denial of HIV status. | Yes | 9 | 4.3 |
|  | No | 194 | 95.7 |

### Qualitative strand

A total of 10 PLWH at SRRH and seven HCPs at CTC participated in IDIs, and we had five FGDs which were conducted and each contained five participants. The study revealed notable differences between clients and HCPs across several demographic characteristics. Clients were generally older (median age 48.5 vs. 27 years) and had lower levels of education, with 80% having only primary education, compared to HCPs, who all held college or university degrees. Women were the majority in both groups, comprising 60% of clients and 71.4% of HCPs. Most participants lived in urban areas, with only one client from a rural setting. Employment status also differed significantly: all clients were self-employed, whereas all HCPs were formally employed in the healthcare sector (Table 4).

**Table 4:** Characteristics of the qualitative study participants who participated in IDIs.

| Characteristic | Category | IDI (Clients) | IDI(HCPs) |
| --- | --- | --- | --- |
| **Age** | Median (IQR) | 48.5(43-52) | 27(26-38) |
| **Sex** | Female | 6(60) | 5(71.4) |
|  | Male | 4(40) | 2(28.6) |
| **Residence** | Rural | 1(10) | 0 |
|  | Urban | 9(90) | 7(100) |
| **Education level** | Illiterate | 0 | 0 |
|  | primary | 8(80) | 0 |
|  | Secondary | 2(20) | 0 |
|  | College & university | 0 | 7(100) |
| **Employment status** | Employed | 0 | 7(100) |
|  | Self-employed | 10(100) | 0 |

### Socio-Demographic Characteristics of FGD Participants

The FGDs included a diverse mix of ages, genders, and experiences with HIV. However, the majority shared similar socioeconomic characteristics: low education levels, self-employment, and long-term HIV diagnoses (Table 5).

**Table 5:**
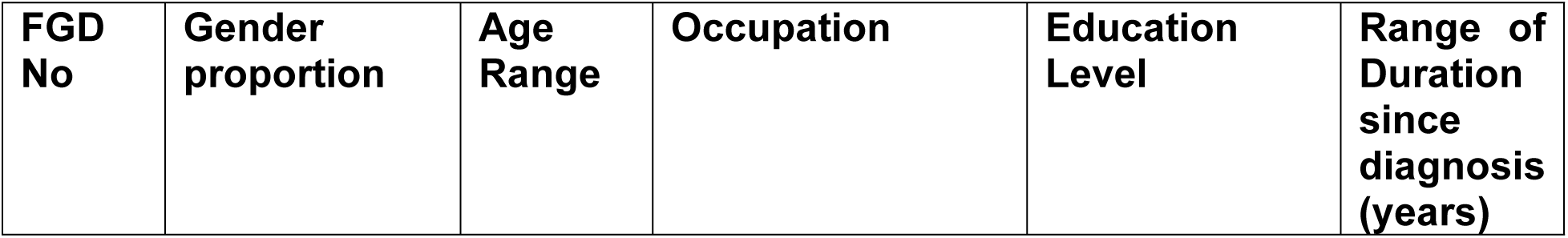
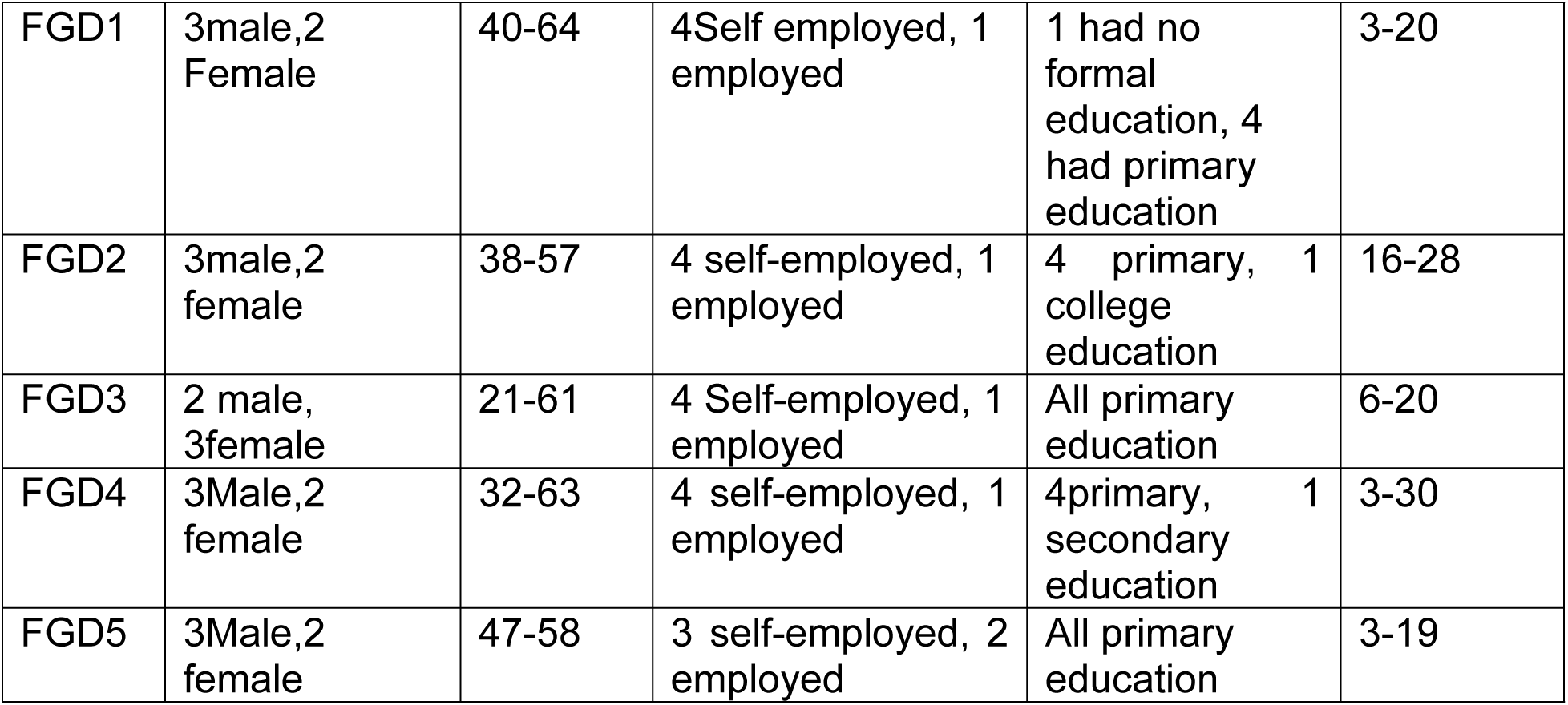
Characteristics of the FGDs study participants (N=25)

### Utilization of APNS

The participants of this study demonstrated varying levels of utilization of APNS, with many attributing their initial exposure to information provided by HCPs, clinic seminars, and external programs such as USAID’s Afya Yangu project. One female PLWH who is also a community healthcare worker stated:

> *“For five years, I worked with the people from the USAID Afya Yangu (My Health) project, and their project also included HIV index testing.”* (FGD2, Female PLWH)

One healthcare provider noted that clients often return to the clinic reporting that they were unable to inform their sexual partners despite their initial intentions to disclose.

> *“It is known that it has its difficulties because many say they will go tell their sexual partners, but when they come back, if you ask them, they say they failed to tell them, so to some extent, it is difficult, and that is why we have been following up continuously.”* (IDI2, Female nurse.)

### Understanding of APNS

Generally, some participants were able to clearly articulate the purpose of APNS, recognizing that it involves identifying and testing all sexual partners, both spouses and casual partners, so they can know their HIV status and access appropriate care or prevention services

> *“It means testing partners and letting them know their HIV status, whether it’s your spouse or even a casual partner… All your sexual partners should know your HIV status and get tested for their health.”* (FGD2, female PLWH)

However, other participants demonstrated only a superficial or limited understanding of the service and reflected on the difficulty they faced in disclosing their HIV status to a seronegative partner, emphasizing that it was only after receiving support and guidance from HCPs that they felt able to take action:

> *“I understood this service a little bit… I had another wife who is not infected, so I had a little difficulty. In the sense of conveying information about my infection status to her. But later, after receiving instructions from health professionals, I took the responsibility of informing my partner.”* (FGD 2, Male PLWH)

### The commonly used APNS Methods

The most commonly used method of APNS reported by participants was the passive referral approach, where the index client personally informs their sexual partners about potential HIV exposure, followed by the provider referral approach. One Female Laboratory technician stated:

> *“Here, mostly we use the method where the index client goes to tell their sexual partners or I, the healthcare provider, go to follow the sexual partner.”* (IDI1, Female Laboratory technician)

### Barriers to APNS

Findings from the qualitative interviews revealed barriers to the uptake of APNS across multiple levels of the SEM.

#### Individual-level barriers

At the individual level, many participants reported a dominant concern about the fear of HIV-related stigma, especially after HIV disclosure, which led some to conceal their HIV status or avoid naming partners. One female PLWH stated:

> *“I was thinking about how society and my partner would perceive me, and how much they would stigmatize me.”* (FGD1, female PLWH)

Participants also expressed fear of IPV, particularly women. They feel uncertain about the other people’s intentions and worry that HIV disclosure through APNS could lead to IPV. As a result, they choose to remain silent rather than risk confrontation or danger. One female PLWH stated:

> *“When you tell someone you’re sick, your heart hesitates first. You know if you announce it directly, they might hit you, harm you. You’re afraid to say something like that, so you just stay quiet, silent.”* (FGD3, Female PLWH)

Additionally, participants noted that multiple or unstable sexual relationships presented significant barriers to effective partner notification, particularly among young women. Moreover, fear of losing financial or material support discouraged disclosure. A male doctor explained:

> *“Young women may be sexually involved with three or even four different people, and they rely on them for financial or other forms of support…they fear that disclosing their status will cause them to lose these “channels” of support.”* (IDI3, Male doctor)

Finally, some HCPs and HIV index testers expressed frustration at the lack of financial support for field visits, noting that they often had to cover transport and related costs themselves. A female nurse stated:

> *“I personally do this work, but have not received payment for a long time.”* (IDI2, Female nurse)

#### Interpersonal-level barriers

Interpersonal dynamics presented significant barriers around APNS. Participants highlighted concerns about disrupting the current relationships, especially when former partners had married or established new families. One female PLWH explained:

> *“I can’t…He’s already married and has his own family, so it would be difficult for me to provide information.”* (FGD1, Female PLWH)

There was gender dynamics as participants emphasized that women often feared backlash, blame, or relationship conflict when initiating HIV disclosure, especially when they were diagnosed before their male partners. In contrast, men were perceived to have more freedom and less vulnerability when disclosing first.

> *“When a woman is identified first and tested, she fails to explain to her husband because she thinks that when I go to tell my partner, either it is her husband or lover, it will seem like she caused it, but for men, it is easier, even if a man tests positive.”* (ID2, Female nurse)

Participants reported a lack of current communication, contact information, or did not know where their former partners resided, making follow-up impossible:

> *“We used to communicate by phone, but later his phone became unreachable… The other one, I’m not sure what the name of his village is.”* (IDI2, Female PLWH)

#### Community-level barriers

At the community level, in some cases, participants reported that traditional practices such as wife inheritance discouraged women from disclosing their HIV status, even when they were aware of the risks to others in the household. The desire to maintain family harmony or avoid disrupting established norms often outweighed health concerns. A female laboratory technician shared her experience:

> *“You’ll find that an older brother has died, and the younger one inherits his wife. Now, she won’t tell the younger brother that “your older brother died with a certain condition and I’m also using certain medication,” she doesn’t say anything.”* (IDI1, Female lab technician)

Participants highlighted significant challenges such as geographical isolation and limited access to healthcare facilities. One male PLWH contrasted the widespread implementation of APNS in urban centers with the persistent gaps in rural communities:

> *“On the other hand, these APNS services have been widely implemented in urban areas, but there is still a problem in rural areas.”* (FGD4, Male PLWH)

Distance to health facilities and associated transportation costs as clients faced challenges in traveling to clinics for testing or follow-up, and HCPs similarly struggled with the financial and logistical burden of conducting outreach visits. One nurse described these difficulties:

> *“Some of our clients come from far, from other districts, so for a person to come from there to test here at the clinic, sometimes it is difficult…sometimes we have to follow up clients, so leaving work to follow a client where they are requires costs, transport issues. So, because some come from far, the cost becomes bigger.”* (IDI2, Female nurse)

#### Structural and institutional barriers

Participants described breakdowns in communication with HCPs, such as unclear explanations about the APNS service or missed opportunities to encourage disclosure. A female PLWH expressed her breakdowns in communication with HCPs as:

> *“If they had told me back then, they would have helped me. But they didn’t. In those days, they didn’t tell you anything. When they called each other, I felt uneasy, like something was wrong.”* (IDI2, Female PLWH)

Several participants noted that inconsistent follow-up from HCPs limited their engagement with the APNS process. One male doctor stated:

> *“We don’t always follow up. We rarely go back to review and say, “Hey, let me call this client and ask if we can do partner notification and test again today.” I think the issue is a lack of structured follow-up for APNS; it’s not fully implemented… We tend to focus more on what happens on the day of the clinic visit.”* (IDI3, Male doctor)

Moreover, healthcare workers themselves acknowledged test kit stockouts as barriers to service provision, and it constrained their ability to conduct both facility-based and community outreach index testing, thereby reducing the reach and efficiency of APNS services. One female nurse reported:

> *“You may need to go test, but find a few supplies. You cannot go to the streets to do mobile testing or Index testing when supplies are few.”* (IDI2, Female nurse)

Several healthcare providers and clients attributed recent declines in the reach and quality of APNS services to the withdrawal of USAID support, which had previously funded outreach and community-based HIV index testing activities. One participant who was a female PLWH expressed community uncertainty and the need for clearer communication:

> *“…After the funding was cut off, especially when that man, pulled out of the WHO, so many of us were shocked. Even though I don’t know much… we need to get clear answers from someone like you, who understands this deeply.”* (FGD2, Female PLWH)

##### Facilitators for APNS

Guided by the SEM, the analysis identified multilevel facilitators influencing the uptake and implementation of APNS. Themes emerged across four levels: individual, interpersonal, community, and organizational.

###### Individual level

At the individual level, participants highlighted that clients’ choices were shaped by their understanding of medical advice from HCPs and supportive counseling from HCPs.

### Medical advice from providers

Clients described how receiving clear, empathetic, and informative advice from HCPs empowered them to initiate disclosure to their partners. This guidance acted as a catalyst for action. A male PLWH stated:

> *“Later, after receiving instructions from health professionals, I took the responsibility of informing my partner.”* (FGD4, Male PLWH)

### Supportive and nonjudgmental counseling from HCPs

Participants emphasized that nonjudgmental and supportive interactions with HCPs played a crucial role in their willingness to engage with and continue using APNS services. One female PLWH captured this sentiment:

> *“If you’re judged, you won’t come back. But if treated with love and care, it gives you courage. Love is powerful.”* (IDI2, Female PLWH)

#### Interpersonal level

This level focused on the direct relationships between clients, their partners, and healthcare providers. Assurance of confidentiality, building rapport, ongoing follow-up, and flexible approaches to partner engagement and team and peer support among HCPs were critical for effective APNS delivery.

### Assurance of confidentiality

One HCP emphasized ethical standards and legal boundaries in maintaining client privacy. This helped build trust and encouraged participation in APNS. A female laboratory technician who was working as the HIV index tester stated:

> *“There are laws and personal ethics. We take an oath never to disclose patient information… And we only inform someone if the tested individual authorizes it.”* (IDI4, Female laboratory technician)

### Building rapport

Establishing rapport with clients was revealed by participants as essential to facilitating HIV disclosure and partner listing. Providers employed empathy and humor to ease the process. One female nurse stated:

> *“Sometimes you really try to sit down with a patient, ask them questions, befriend them, make them laugh, just so that they can open up and name someone.”* (IDI1, Female nurse)

### Flexible approaches to partner engagement

Providers described adapting APNS strategies to suit individual needs, demonstrating flexibility in how partners were reached. This included direct, in-person contact when phone communication was unreliable. One female client explained the value of face-to-face engagement:

> *“You’d have to go in person. Calling might not work; he might change numbers. If you go and talk face-to-face, he might accept.”* (IDI4, Female PLWH)

Similarly, a provider described strategies to create a supportive environment at the clinic for partner testing without revealing identities:

> *“We help them, or they can bring their partner even here at the clinic to come test together… we pretend as if we don’t know them… this reduces the pressure.”* (IDI2, Female nurse)

### Ongoing follow-up

HCPs emphasized the importance of ongoing follow-up as a key strategy to ensure continuity and encourage clients to use APNS. One nurse described the practice of providing medication with a timeframe that motivates timely partner engagement and notification:

> *“We always follow up… we give them medicine for fourteen days, but we ask them if it is possible to bring their sexual partners within seven days.”* (IDI2, Female Nurse)

### Team and peer support among providers

HCPs highlighted the importance of collaboration when encountering difficulties during APNS counseling sessions. With client consent, they often consulted colleagues to provide additional support and ensure comprehensive care. One nurse described this collaborative approach:

> *“You might talk with the client and realize you have reached a limit… so you find a colleague counselor and you assist each other.”* (IDI2, Female Nurse)

#### Community level

At the community level, the focus was on community accessibility and delivery of services in social spaces, including home-based and mobile testing.

### Community accessibility

Providers actively engaged in proactive outreach by offering APNS and HIV testing services directly within communities or following up with clients at their locations. These approaches improved accessibility and convenience while helping to reduce the stigma associated with facility-based testing. A female lab technician explained:

> *“If a person wants us to follow them in the community, we follow them and test them there.”* (IDI1, Female laboratory technician)

#### Organizational level

Organizational-level factors included staff training and incentives that reinforced client participation in APNS and motivated providers.

### Training and refresher education

Ongoing training for both HCPs and PLWH was reported to enhance understanding of APNS and its benefits. A female PLWH expressed how training inspired them to encourage partner testing:

> *“They teach us about partner notification. This motivates us to talk to our partners and encourage them to get tested.”* (FGD2, Female PLWH)

### Incentives for participation

Participants emphasized that health facilities to use simple incentives to motivate clients to disclose their HIV status and bring their partners for HIV testing. They insisted that PLWH who arrived with their partners be given a priority service as a way to encourage others to do the same. One participant shared:

> *“The one who comes, even if late, if they come with their partner, we serve [them] first… to motivate the other.”* (IDI2, Female nurse)

The summary below outlines the key barriers and facilitators influencing the utilization of APNS as reported by study participants (Fig. 3).

**Fig 3:**
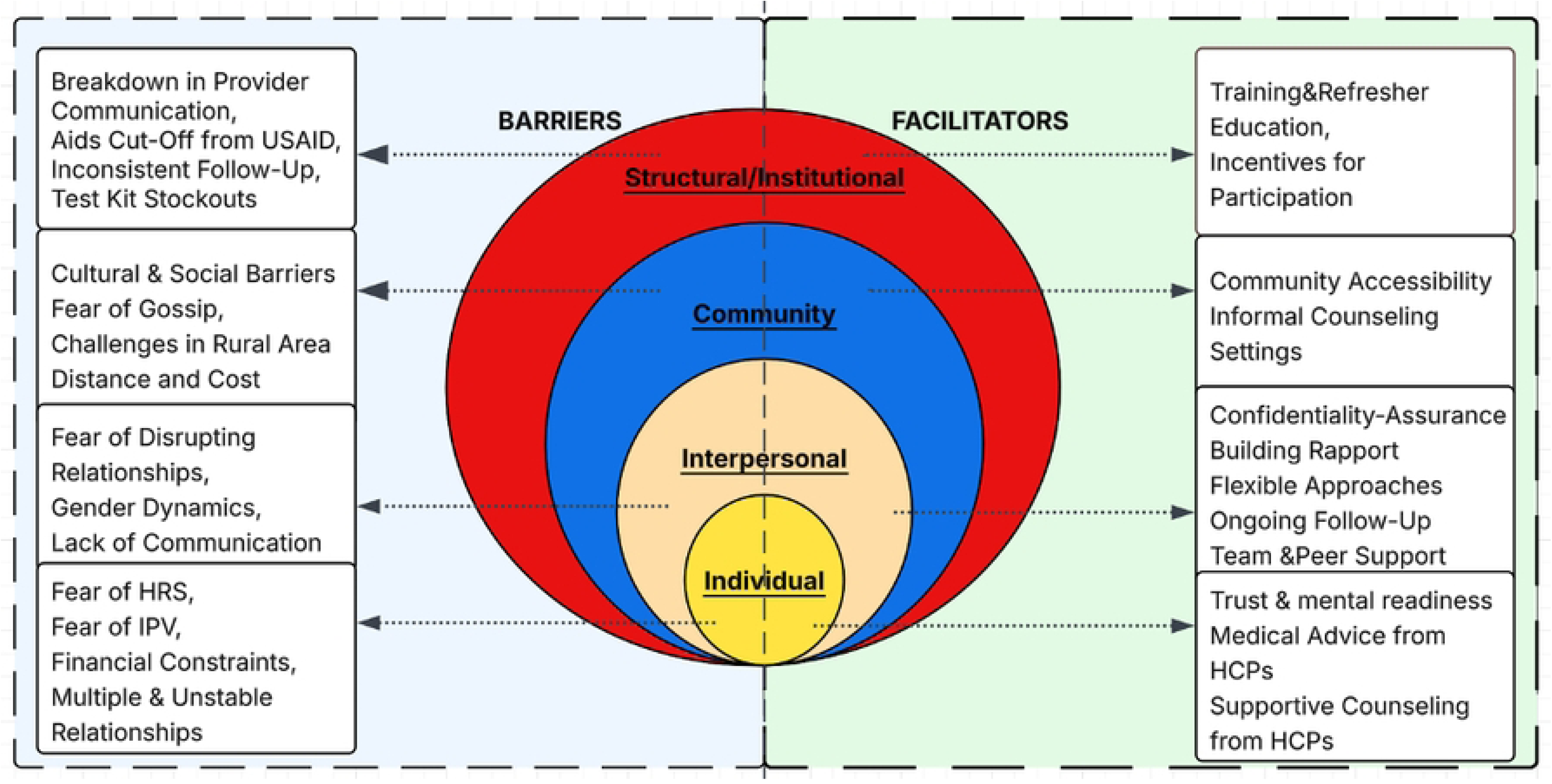
Barriers and Facilitators Influencing the Use of APNS Across the Socioecological Model.

## Discussion

In this study, quantitative findings revealed that close to half 40% of study participants had used APNS to notify their sexual partner about the potential exposure to HIV infection because of their HIV seropositive status, underscoring the limited uptake of this essential HIV prevention strategy. This was further supported by qualitative findings, which also revealed that although many participants expressed willingness to disclose their HIV status to sexual partners as part of the APNS process, in practice, HIV disclosure to sexual partners through APNS remains a significant challenge. This limited utilization of APNS among the participants contrast with that reported in regional studies where utilization of APNS is higher; in Kenya (69%), Ethiopia (65.5%), Eastern Tanzania (56%), and Northern Tanzania (62%) [15, 18–20]. Contextual differences may explain this discrepancy due to differences in health system structure, client education, or provider engagement strategies in those areas.

Our findings confirmed that a substantial proportion (86%) of hypertensive PLWH were hypertensive but unaware of being hypertensive, suggesting underdiagnosis and missed opportunities for integrated care among PLWH. That is consistent with Kanyike et al (2024) study, which was conducted in Uganda, which revealed that the majority of 68% of PLWH were unaware of their hypertensive condition [42]. These results call for expansion of integrated services that address both chronic comorbidities and sexual health which could help capture missed populations and provide more holistic support.

### Knowledge and mental health as key determinants of APNS uptake

Our findings demonstrate that higher HIV disclosure knowledge significantly increased the likelihood of APNS use. Participants with moderate and high knowledge had nearly two to three times greater odds of utilizing APNS, respectively. When PLWH understand the purpose, benefits, and process of disclosing their HIV status and notifying partners, they are more confident and willing to participate in APNS. That is similar to the studies, which were conducted in Uganda and Ethiopia by Klabbers et al. (2021) and Oljira et al. (2024), respectively, which revealed that clients with a better understanding or knowledge of HIV transmission and the APNS process were significantly more likely to notify partners [18, 43]. These findings were reinforced by our qualitative interviews, where PLWH with more exposure to health talks and provider counseling expressed greater trust in the system and described feeling “empowered” to notify partners through APNS. Participants who had limited information about APNS reported confusion, fear, and reluctance to participate. This alignment underscores the importance of continuous education and personalized counseling in enhancing APNS uptake.

Depression limits APNS uptake, with each unit increase in depression score reducing APNS engagement by 5%. This trend is consistent with the study from Ethiopia, which revealed that having any degree of depression decreased the probability of using APNS by 88% compared with those free of depression [18]. Qualitative findings further contextualized this relationship; several participants described emotional distress, hopelessness, and social isolation following diagnosis, which limited their motivation or capacity to engage in APNS. These findings highlight the necessity of embedding psychosocial support and mental health education within HIV care. Providers and peer educators should be trained to recognize depressive symptoms and provide HIV disclosure counseling that boosts self-efficacy and addresses emotional well-being.

### Barriers to APNS: multi-level and structural

This study identified multiple barriers to the utilization of APNS. Quantitatively, the most commonly reported barrier was fear of stigma (47.8%), followed by fear of embarrassment (30.4%) and fear of losing emotional support or autonomy (28.3%). This is almost consistent with Chegoloi et al (2021) study, which revealed that nearly half of the respondents reported both fear of stigma, fear of embarrassment, and 24.3% of respondents had fear of losing emotional support or autonomy [9]. The convergence between our findings and prior research underscores the enduring role of HIV-related stigma as a structural and interpersonal barrier. Qualitative findings in this study further revealed how fear of blame and social rejection discouraged HIV disclosure and participation in APNS, particularly among women who depended on partners for social or financial support. This reflects the social-ecological interplay between individual fears and broader gender and community dynamics. Interventions to improve APNS uptake should therefore move beyond individual counseling to incorporate stigma-reduction strategies at community and health-system levels, including strengthening confidentiality mechanisms, promoting positive messaging about disclosure, and addressing gendered power imbalances that heighten vulnerability to stigma and blame.

Moreover, confidentiality emerged as a key barrier to the utilization of APNS in both quantitative and qualitative findings. This study quantitatively identified a smaller proportion of participants who reported concerns about confidentiality 13%, yet qualitative narratives revealed that fears of disclosure and mistrust in information handling remained significant deterrents. That is inconsistent with another study by Goyette et al (2016), which revealed that 50% of clients who declined to participate in APNS worried about a breach of confidentiality [25]. This discrepancy may reflect contextual differences in healthcare systems, the level of trust in service providers, or the effectiveness of confidentiality safeguards communicated to clients during APNS counseling.

### Facilitators: trust, tailored engagement, and structural support

This study revealed that confidentiality and building rapport between clients and HCPs were vital in facilitating APNS. That is similar to the Liu et al. (2022) study, which was conducted in China, and it identified facilitators for successful APNS implementation, including building rapport and assuring confidentiality [44]. The same facilitators were reported in the systematic review, which was conducted by Tayakoli et al (2024) [45]. Together, these findings emphasize that APNS effectiveness depends not only on structural availability but also on the quality of interpersonal relationships within healthcare delivery. Strengthening provider communication skills, and ensuring consistent confidentiality safeguards can therefore enhance client engagement and sustain APNS outcomes.

Community- and system-level facilitators included home visits and ongoing staff training, all of which contributed to a more supportive HIV disclosure environment through APNS. In the study by Liu et al. (2022), it was identified that home visits and ongoing staff training contribute to a more supportive HIV disclosure environment [44]. These enablers reflect global evidence supporting patient-centered HIV service models that reduce access barriers and normalize APNS. The study findings reaffirm that a well-functioning, empathetic health system is essential for effective APNS implementation. Policies should support a multi-level approach to APNS by promoting client-centered counseling, strengthening provider-client trust through training and confidentiality safeguards, and expanding community-based outreach and digital notification tools to create a supportive environment for HIV disclosure.

### Broad limitations

The study results may not be generalizable to all people with HIV infection in the entire community population and to rural areas because it was conducted at the single health facility and in a single urban region. Moreover, the present study has some other limitations, such as the utilization of APNS was assessed through self-reporting questions, which may overestimate or underestimate the actual prevalence due to social desirability or recall bias. Additionally, our study might have suffered from a recall bias due to a time lag. The study employed a cross-sectional study design, which made it difficult to establish a causal relationship between the outcome and the predicting variables. Lastly, the cross-sectional nature of the quantitative strand precludes causal inference.

### Broader implications

The insights from this study offer valuable guidance for APNS policy and programming globally, especially in resource-limited, high-HIV-burden contexts. First, the findings reinforce that public health strategies regarding APNS must be tailored to community realities, incorporating gender, age, social stigma, and mental health into HIV prevention planning. Second, the evidence affirms the importance of health system resilience and provider capacity, especially where donor funding fluctuates. Without institutional commitment to APNS training, supplies, and supervision, implementation will remain uneven and unsustainable. Finally, this study highlights the potential of participatory approaches that value the lived experiences of PLWH. Their recommendations, spanning digital innovations, community outreach, and dignified care, should shape future APNS strategies. As countries strive toward the UNAIDS 95-95-95 targets, optimizing APNS through grounded, multi-sectoral approaches will be essential to reaching undiagnosed individuals and curbing new infections.

### Conclusion

This mixed-methods study provides critical insights into the multifaceted barriers and facilitators influencing the APNS among PLWH in Tanzania. Despite the proven public health value of APNS, its utilization remains limited in the study area, particularly due to stigma, mental health challenges, gendered power dynamics, and systemic health system constraints. However, the findings also reveal promising enablers, including knowledge on HIV disclosure, trust in providers, emotional readiness, community outreach, and digital tools, that can be leveraged to improve APNS service delivery. The study highlights the importance of integrating APNS within a comprehensive, client-centered framework that addresses mental health, gender equity, hypertension screening, and systemic weaknesses in the healthcare system. To effectively scale up APNS and reduce HIV transmission, policymakers, healthcare providers, and researchers must collaborate to design context-specific, inclusive, and sustainable models that respond to the diverse needs and lived realities of those affected.

Our findings underscore that APNS uptake is shaped by cognitive, emotional, social, and structural dynamics. Increasing HIV disclosure knowledge, addressing mental health concerns, and fostering strong client-provider relationships are essential strategies for improving participation in APNS. We recommend the importance of integrated counseling, provider capacity building, and community education. Multi-level, context-specific approaches, particularly those that are gender-sensitive and responsive to the needs of older adults and high-risk populations, are critical. Future research should explore the effectiveness, sustainability, and cost-effectiveness of tailored APNS interventions, and also examine how to incorporate APNS as a strategy to enhance PrEP screening and uptake, including the integration of HIV care with chronic disease management. Collectively, these efforts are essential to ensure APNS reaches its full potential as a cornerstone of HIV prevention and care in Tanzania and similar settings.

## Data Availability

Data cannot be shared publicly because of ethical restrictions related to the confidentiality of research participants living with HIV. The data contains potentially identifying and sensitive participant information. Data are available from the Kilimanjaro Christian Medical University College (KCMUCo) Ethical Committee and the National Institute for Medical Research (NIMR), Tanzania, for researchers who meet the criteria for access to confidential data.

## Acknowledgement

I thank Almighty God for His guidance, strength, and blessings throughout this journey. I would also like to express my deepest gratitude to Prof. Blandina Mmbaga, Prof. John Bartlett, Prof. Charles Muiruri, and Dr. Florida Muro for their invaluable guidance and support throughout this project. I am sincerely thankful to the Forgaty International Center for funding this study as one of the pilot grants. My heartfelt appreciation goes to the participants from SRRH, whose willingness and openness made this study possible. Finally, I wish to acknowledge the administration of SRRH and the CTC team for their essential contributions to the successful completion of this work.

## Supporting Information

**S1 File. Quantitative-research-questionnaire**

**S1 Fig. Illustration of the Social Ecological Model (SEM).**

**S2 Fig. Pie chart showing the distribution of types of APNS**

**S3 Fig. Barriers and Facilitators Influencing the Use of APNS Across the Socioecological Model**

## Notes

### Competing Interest Statement

The authors have declared no competing interest.

### Author Declarations

The study was approved by KCMC University ethical committee (No. 2664) and the National Institute for Medical Research (NIMR) (No. NIMR/HQ/R.8a/Vol.IX/4866). Written informed consent was obtained from the study respondents individually. The right of the respondent to withdraw from the interview or not to participate at all was assured. To maintain participant confidentiality, the interviews were conducted in a private room with the door closed, and all of the documents with participants' information were well protected in a locked cabinet only the principal investigator had access to the key.

